# Anakinra and Intravenous IgG versus Tocilizumab in the Treatment of COVID-19 Pneumonia

**DOI:** 10.1101/2020.09.11.20192401

**Authors:** Massa Zantah, Eduardo Dominguez Castillo, Andrew J. Gangemi, Maulin Patel, Junad Chowdhury, Steven Verga, Osheen Abramian, Matthew Zheng, Kevin Lu, Arthur Lau, Justin Levinson, Hauquing Zhao, Gerard J. Criner, Roberto Caricchio, for the Temple University COVID-19 Research Group

## Abstract

**Background:** COVID-19 can lead to acute respiratory failure and an exaggerated inflammatory response. Studies have suggested promising outcomes using monoclonal antibodies targeting IL-1β (Anakinra) or IL6 (Tocilizumab), however no head to head comparison was done between the two treatments. Herein, we report our experience in treating COVID-19 pneumonia associated with cytokine storm with either subcutaneous Anakinra given concomitantly with intravenous immunoglobulin (IVIG), or intravenous Tocilizumab.

**Methods:** Comprehensive clinical and laboratory data from patients with COVID-19 pneumonia admitted at our hospital between March and May 2020 were collected. Patients who received either Anakinra/ IVIG or Tocilizumab were selected. Baseline characteristics including oxygen therapy, respiratory status evaluation using ROX index, clinical assessment using NEWS score and laboratory data were collected. Outcomes included mortality, intubation, ICU admission and length of stay. In addition, we compared the change in ROX index, NEWS score and inflammatory markers at days 7 and 14 post initiation of therapy.

**Results:** 84 consecutive patients who received either treatment (51 in the Anakinra/ IVIG group and 33 in the Tocilizumab group) were retrospectively studied. Baseline inflammatory markers were similar in both groups. There was no significant difference regarding to death (21.6% vs 15.2%, p 0.464), intubation (15.7% vs 24.2%, p 0.329), ICU need (57.1% vs 48.5%, p 0.475) or length of stay (13+9.6 vs 14.9+11.6, p 0.512) in the Anakinra/IVIG and Tocilizumab, respectively. Additionally, the rate of improvement in ROX index, NEWS score and inflammatory markers was similar in both groups at days 7 and 14. Furthermore, there was no difference in the incidence of superinfection in both groups.

**Conclusion:** Treating COVID-19 pneumonia associated with cytokine storm features with either subcutaneous Anakinra/IVIG or intravenous Tocilizumab is associated with improved clinical outcomes in most subjects. The choice of treatment does not appear to affect morbidity or mortality. Randomized controlled trials are needed to confirm our study findings.

**Funding:** None.

## Introduction

Since the first case was reported in December 2019, the severe acute respiratory syndrome coronavirus 2 (SARS-CoV-2) pandemic has resulted in over 10 million confirmed infections and over 500,000 deaths worldwide^1,2^ The number of cases continues to rise in many countries, including in the US. The disease has overwhelmed the health care systems globally and nationally. Thus far, no effective therapy has been proven to treat COVID-19 disease. Available therapies include supportive care, antibiotics and invasive and noninvasive oxygen support. In addition, off-label therapies including antiretrovirals, antiparasitic agents, anti-inflammatory medications, and convalescent plasma have been used.^3-8^

In its severe form, the virus can lead to a life-threatening pneumonia and acute respiratory distress syndrome (ARDS). Although the mechanisms of COVID-19-induced lung injury are still being examined, cytokine storm has been thought to play a role in disease pathophysiology. This form of hyperactive and dysregulated immune response may lead to hyperinflammatory form of ARDS, extrapulmonary multiple-organ failure, and is associated with critical illness and increased mortality.^9-11^ Cytokine storm-related respiratory failure carries significant morbidity; in a large retrospective review of hemophagocytic lymphohistiocytosis (HLH), a form of “cytokine storm syndromes,” 66% of patients required advanced mechanical ventilatory support, with 40-50% requiring additional vasopressor and renal replacement therapy; mortality approached 53%.^12^ Some of the proposed treatments for COVID-19 pneumonia aim to target the aforementioned inflammatory cascade, monoclonal antibodies targeting interleukins (e.g. Sarilumab, Anakinra, and Tocilizumab) are still under investigation in large randomized clinical trials (NCT04322773, NCT04315298) however evidence from retrospective, observational studies and case reports suggest the effectiveness of some of these therapies. Herein, we present our experience in the treatment of COVID-19 disease, in which we used both recombinant IL-1 receptor antagonist and IL6 monoclonal antibody targeting agents and compared them in our population of COVID 19 patients with moderate to severe hypoxemic repsiroatry failure due to pneumonia. These therapies were given in addition to a corticosteroid, which has been recently found to significantly reduce mortality in patients with COVID-19 on respiratory support. Furthermore, we present an approach using intravenous immunoglobulin (IVIG) in conjunction with subcutaneous anakinra. The combination of IVIG, anakinra, and corticosteroids has been used with success in treating the immune dysregulation associated with secondary HLH and macrophage activation syndrome.^13–15^

## Material and Methods

### Study Design and Population

This is a single center retrospective review conducted at Temple University Hospital. We included consecutive patients admitted between March 17th and May 7th, 2020 and treated for COVID-19 viral pneumonia. The diagnosis of COVID-19 pneumonia was based on a constellation of clinical, radiographic and laboratory findings. Clinical criteria include signs or symptoms of COVID-19 disease such as fever, cough or shortness of breath. All patients had computed tomography (CT) of the chest on admission to detect and quantitate the extent of pulmonary infiltrate. The presence of bilateral peripheral ground-glass opacities (GGOs) was considered highly suggestive of COVID-19 pneumonia. Laboratory findings that help make the diagnosis include positive nasopharyngeal polymerase chain reaction (PCR) for SARS-CoV-2 and/or lymphopenia, elevated inflammatory markers including C-reactive protein (CRP), lactate dehydrogenase (LDH), Ferritin, and D dimer. Patients who received Anakinra with intravenous immunoglobulin (IVIG), or Tocilizumab were included in the study.

### Study Treatments

The decision for the treatment was made after a multidisciplinary discussion that involved pulmonary and rheumatology specialists^16^ A consensus was based on: 1) worsening respiratory status defined as increased oxygen supplementation required to maintain SpO_2_≥ 93%, and 2) elevation above 3-fold the upper normal level of at least two of the following markers: CRP, ferritin, D-dimer, LDH and cardiac troponin. Patients meeting the above criteria were treated with either therapy unless contraindicated. Anakinra was given subcutaneously at a dose of 100mg every 6 hours or 100mg every 12 hours for renal impairment for 7 days; along with IVIG 0.5g/kg/day for 3 days. Tocilizumab was given intravenously and dosed 8 mg/kg in a single infusion and repeated within 12 hrs if conditions worsened or at 24 hrs if no clinical improvement.

### Data Collection and Measures

All the data were collected using our electronic medical records. Demographics included age, gender, ethnicity, and body mass index (BMI).

Comorbidities including hypertension (HTN) diabetes (DM), coronary artery disease (CAD), congestive heart failure (CHF), chronic kidney disease (CKD), dialysis-dependent end-stage renal disease (ESRD), human immunodeficiency status (HIV), and history of malignancy were gathered for all patients.

Baseline laboratory data included inflammatory markers (CRP, LDH, Ferritin, D dimer, Fibrinogen, Triglyceride), absolute lymphocyte count, Albumin, kidney function measured by blood urea nitrogen/creatinine (BUN/Cr) ratio, liver function testing; Alanine transaminase and Aspartate transaminase (ALT and AST).

Respiratory function status was evaluated using the ROX index. This is defined as the ratio of oxygen saturation as measured by pulse oximetry/FIO2 to respiratory rate. ROX index has been assessed as a predictor of high-flow nasal cannula treatment and need for intubation. Higher scores (ROX ≥4.88) predict low risk for intubation.

Clinical assessment for patients was detected using the National Early Warning Scores 2 (NEWS2). The NEWS2 score was developed by the Royal College of Physicians and modified for use with hypoxic and hypercapnic respiratory failure; it was also validated in a small study of COVID-19 patients. It takes into account six physiological findings and one observation (respiratory rate, oxygen saturation, supplemental oxygen status, temperature, systolic blood pressure, heart rate and level of consciousness). A score of ≥5 signifies a need for ICU monitoring.^17,18^

### Clinical Outcomes

Our co-primary endpoints were death and need for intubation. Secondary outcomes included the need for an intensive care unit (ICU), hospital length of stay (LOS), changes in ROX index, NEWS2 score, and laboratory data at day 7 and 14 from initiation of treatment. Additionally, we reported secondary infections (bacteremia and/or ventilator associated pneumonia) in both groups.

### Data Analysis

Data are presented as means with standard deviations (SD) for continuous variables or as percentages for categorical variables. We used a chi-squared test for categorical data, and a two-sided t-test for continuous data. P values < 0.05 were considered statistically significant. The Fisher exact test was used for simple between-group comparisons. Survival data was presented on Kaplan Meier curve. The software we used to run the statistics was Stata 14 *(Stata Statistical Software: Release 14*. College Station, TX: StataCorp LP). A univariate subanalysis was performed to compare the results between dead and survivors of the study population. The Temple University Hospital Institutional Review Board approved the protocol.

## Results

### Baseline characteristics

A total of 51 patients who were treated with subcutaneous Anakinra and IVIG and 33 patients who received intravenous Tocilizumab were identified for the study. Their baseline characteristics including demographics and comorbidities are listed in Table-1. The majority of patients in both groups were males (63.1%), with a mean age of 60.6±13.4 years. Mean BMI was 31.4±8.0 kg/m^2^. The diagnosis of the majority of patient was made using nasopharyngeal swab for Cov2-SAR PCR (77.4%). There was no significant difference in regard to comorbidities in both groups, including COPD, asthma, DM, HTN, CAD, CHF, CKD and ESRD. Unless contraindicated, all patients requiring oxygen therapy received Corticosteroids, with a mean dose (methylprednisolone equivalent) of 218.1±149.7 mg in the Anakinra/IVIG versus 246.2±129.8 mg in the Tocilizumab group, p= 0.37. In both groups, patients had similarly elevated baseline inflammatory markers including CRP (8.5±5.9 versus 9.4±8.6 mg/L), LDH (508.1±575.6 versus 345.1±106.4 U/L), D dimer (6126.1±17554.9 versus 1398.4±2798.8 ng/mL), fibrinogen (508.5±117.8 versus 537.6±138.2 mg/dL), and triglycerides (137.4±82.6 versus 146.5±96.8 mg/dL), in the Anakinra/IVIG and Tocilizumab respectively (Table-2).

**Table 1-.** Baseline Characteristics:

| Demographics | Total<br>n=84 | Anakinra/IVIG<br>n=51 | Tocilizumab<br>n=33 | P-value |
| --- | --- | --- | --- | --- |
| Gender (%) |  |  |  | 0.704 |
| Male | 53 (63.1) | 33 (64.7) | 20 (60.6) |  |
| Female | 31 (36.9) | 18 (35.3) | 13 (39.4) |  |
| Mean Age, years | 60.6±13.4 | 62.7±12.3 | 57.4±14.6 | 0.074 |
| Ethnicity (%) |  |  |  | 0.660 |
| AA | 45 (53.6) | 28 (54.9) | 17 (51.5) |  |
| Hispanic | 25 (29.8) | 15 (29.4) | 10 (30.3) |  |
| Caucasian | 13 (15.5) | 8 (15.7) | 5 (15.2) |  |
| Not known /NON<br>Hispanic | 1 (1.2) | 0 (0.0) | 1 (3.0) |  |
| Mean BMI (kg/m<sup>2</sup>) | 31.4±8.0 | 30.0±7.7 | 33.7±8.1 | 0.035 |
| COVID PCR<br>positive status (%) | 65 (77.4) | 41 (80.4) | 24 (72.7) | 0.412 |
| **Comorbidities** |  |  |  |  |
| COPD (%) | 23 (27.4) | 17 (33.3) | 6 (18.2) | 0.128 |
| Asthma (%) | 8 (9.5) | 4 (7.8) | 4 (12.1) | 0.514 |
| HTN (%) | 60 (71.4) | 37 (72.5) | 23 (69.7) | 0.777 |
| DM (%) | 36 (42.9) | 18 (35.3) | 18 (54.5) | 0.082 |
| CAD (%) | 16 (19.0) | 11 (21.6) | 5 (15.2) | 0.464 |
| CHF (%) | 17 (20.2) | 8 (15.7) | 9 (27.3) | 0.197 |
| CKD (%) | 12 (14.3) | 8 (15.7) | 4 (12.1) | 0.648 |
| ESRD on dialysis (%) | 11 (13.1) | 8 (15.7) | 4 (9.1) | 0.382 |
| Malignancy (%) | 4 (4.8) | 4 (7.8) | 0 (0.0) | 0.099 |
| HIV (%) | 4 (4.8) | 1 (2.0) | 3 (9.1) | 0.134 |
| Highest Corticosteroid dose (Methylprednisolone) (mg) | 229.1±142.1 | 218.1±149.7 | 246.2±129.8 | 0.379 |

**Table-2.** Baseline Laboratory, Respiratory Function and Clinical Data

|  | **Total**<br>**n=84** | **Anakinra/IVIG**<br>**n=51** | **Tocilizumab**<br>**n=33** | **P value** |
| --- | --- | --- | --- | --- |
| **Baseline laboratory Inflammatory Markers** |  |  |  |  |
| CRP (mg/L) | 8.9±7.0 | 8.5±5.9 | 9.4±8.6 | 0.622 |
| Ferritin (ng/mL) |  |  |  | 0.081 |
| Mean | 1599.2±5084.2 | 2276.8±6413.5 | 520.2±434.2 |  |
| Median (IQR) | 563.5 (247.0-993.0) | 628.0 (360.0-1090.0) | 348.0 (175.0-878.0) |  |
| LDH (U/L) | 446.4±463.4 | 508.1±575.6 | 345.1±106.4 | 0.068 |
| AST (U/L) |  |  |  | 0.153 |
| Mean | 116.8±386.5 | 156.8±484.8 | 51.2±71.1 |  |
| Median (IQR) | 37.5 (25.0-66.0) | 39.0 (24.0-80.0) | 36.0 (30.0-47.5) |  |
| ALT (U/L) |  |  |  | 0.461 |
| Mean | 97.1±25.5 | 111.7±297.0 | 73.2±147.5 |  |
| Median (IQR) | 43.0 (22.0-69.0) | 41.0 (22.0-74.0) | 43.0 (21.5-59.5) |  |
| Albumin (g/dL) | 2.6±0.4 | 2.5±0.4 | 2.6±0.4 | 0.655 |
| D-dimer (ng/mL) |  |  |  | 0.087 |
| Mean | 4328.3±14055.6 | 6126.1±17554.9 | 1398.4±2798.8 |  |
| Median (IQR) | 843.0 (521.0-1745.0) | 1073.5 (606.5-2669.5) | 789.0 (387.0-909.0) |  |
| Fibrinogen (mg/dL) | 519.3±163.7 | 508.5±177.8 | 537.6±138.2 | 0.485 |
| Triglycerides | 141±87.8 | 137.4±82.6 | 146.5±96.8 | 0.691 |
| (mg/dL) |  |  |  |  |
| Absolute Lymphocyte Count (K/mm<sup>3</sup>) | 0.9±0.6 | 0.9±0.6 | 0.9±0.6 | 0.838 |
| BUN/Creatinine ratio | 23.9±12.4 | 23.9±13.4 | 23.9±10.8 | 0.989 |
| **Respiratory Function** |  |  |  |  |
| Mean ROX index at day 0 | 11.6±7.5 | 13.6±7.6 | 8.4±6.3 | 0.002 |
| Maximum Oxygen Support |  |  |  | 0.191 |
| None (%) | 6 (7.1) | 5 (9.8) | 1 (3.0) |  |
| NC (%) | 38 (45.2) | 27 (52.9) | 11 (33.3) |  |
| HFNT (%) | 20 (23.8) | 9 (17.6) | 11 (33.3) |  |
| NIPPV (%) | 3 (3.6) | 2 (3.9) | 1 (3.0) |  |
| IMV (%) | 16 (19.0) | 8 (15.7) | 8 (24.2) |  |
| ECMO (%) | 1 (1.2) | 0 (0.0) | 1 (3.0) |  |
| **Clinical Data** |  |  |  |  |
| NEWS score on day 0 | 6.3±3.6 | 5.6±3.5 | 7.5±3.5 | 0.017 |

\*NC- Nasal cannula, HFNT- High flow nasal Therapy, NIPPV- Non-invasive Positive Pressure Ventilation, IMV - invasive mechanical Ventilation, ECMO- Extracorporeal Membrane oxygenation, NEWS - National early warning Score

In regard to clinical assessment, patients in the Tocilizumab group had worse baseline respiratory function and NEWS score. ROX index was 13.6±7.6 versus 8.4±6.3, p 0.002 and NEWS score was 5.6±3.5 versus 7.5±3.5, p 0.017 in the Anakinra/ IVIG compared to Tocilizumab. The majority of patients in both groups were on low-flow nasal cannula, however, although not statistically significant, more patients in the Tocilizumab group required high-flow nasal cannula (33.3% versus 17.6%) and invasive mechanical ventilation (24.2% versus 15.7%, p=0.191) compared to the Anakinra/IVIG group (Table-2).

### Clinical Outcomes

There was no significant difference in mortality among patients who received Anakinra/IVIG or Tocilizumab (21.6% versus 15.2%, p 0.464. Figure-1). Furthermore, there was no significant difference in regard to other clinical outcomes including intubation (15.7% versus 24.2%, p 0.329), need for ICU (57.1% versus 48.5%, p=0.475), length of stay (13.4±9.6 versus 14.9±11.6, p=0.512) for Anakinra/IVIG compared to Tocilizumab respectively. Although sicker at baseline, patients in the Tocilizumab group when compared to patients in the Anakinra/IVIG had the same rate of improvement in clinical parameters including change in ROX index from baseline at day 7 (6.6±6.5 versus 4.9±6.6, p 0.257) and day 14 (7.8±4.8 versus 6.5±5.8, p 0.537). Similar results were noted regarding change in NEWS score at day 7 (−2.1±3.7 versus −2.2±3.1, p 0.962) and day 14 (−2.2±4.0 versus −2.2±2.0, p 0.989) in the Tocilizumab versus Anakinra/IVIG. We found similar rates of secondary infections (17.6% versus 24.2%, p 0.462) in both groups (Table-3).

**Figure 1.**
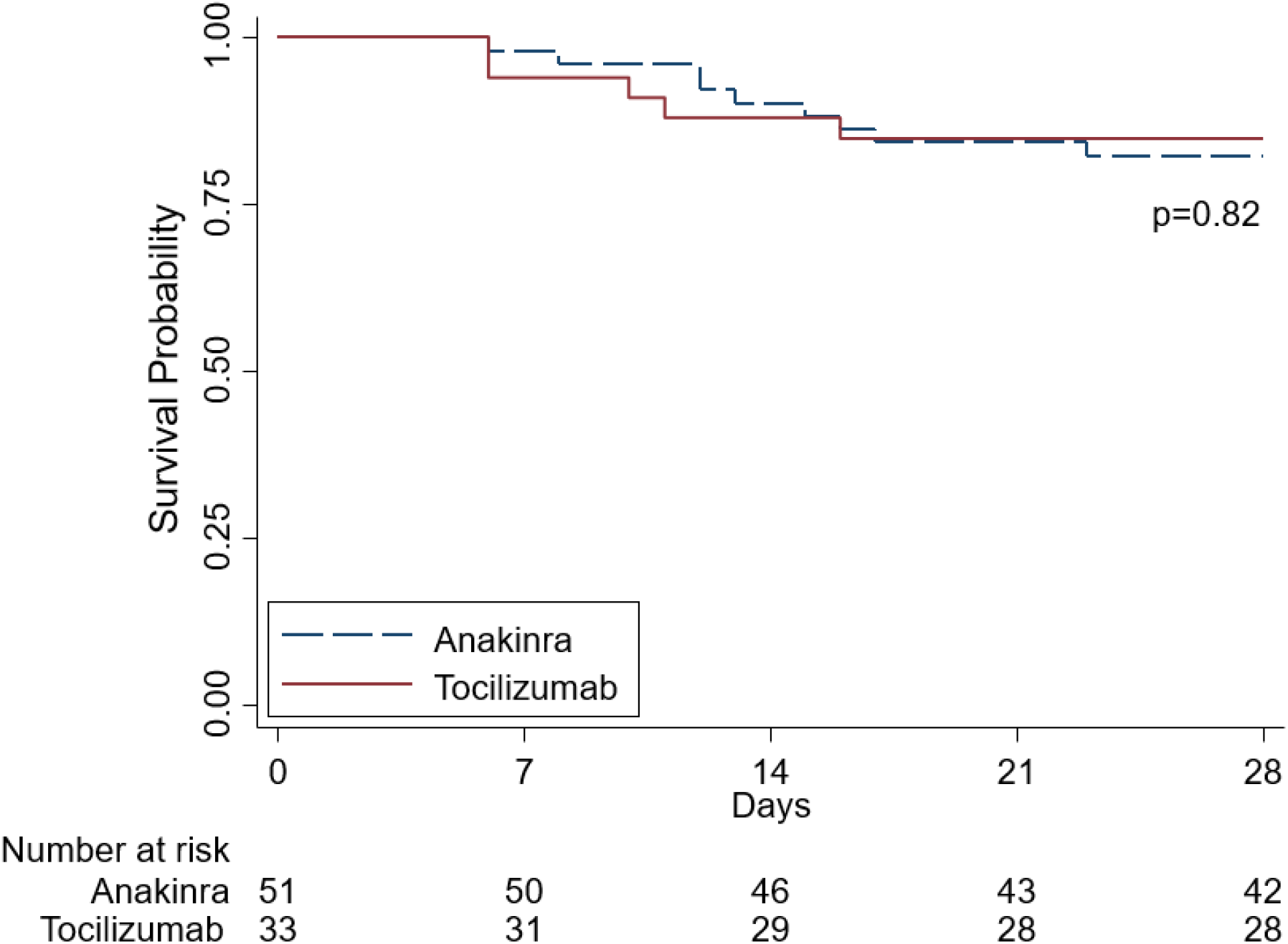
Survival Probability displaying no difference in survival between patients treated with Anakinra/IVIG vs. Tocilizumab.

**Table-3.** Clinical Outcomes

| **Outcome** | **Total**<br>**n=84** | **Anakinra/IVIG**<br>**n=51** | **Tocilizumab**<br>**n=33** | **P-value** |
| --- | --- | --- | --- | --- |
| Death (%) | 16 (19.0) | 11 (21.6) | 5 (15.2) | 0.464 |
| Intubation (%) | 16 (19.0) | 8 (15.7) | 8 (24.2) | 0.329 |
| Need for ICU (%) | 36 (52.9) | 20 (57.1) | 16 (48.5) | 0.475 |
| Composite outcome<br>(death, intubation and<br>need for ICU) (%) | 38 (45.2) | 21 (41.2) | 17 (51.5) | 0.352 |
| Hospital Length of<br>Stay (LOS) (days) | 14±10.4 | 13.4±9.6 | 14.9±11.6 | 0.512 |
| ICU LOS (days) |  |  |  | 0.474 |
| Mean | 8.5±7.0 | 7.7±4.5 | 9.5±9.3 |  |
| Median (IQR) | 6.5 (3.0-12.0) | 6.5 (4.0-12.0) | 7.5 (2.5-11.5) |  |
| Duration of Invasive Mechanical of Ventilation (days) | 13.3±10.9 | 12.9±13.2 | 13.6±9.6 | 0.904 |
| Change in NEWS score from baseline at day 7 | -2.1±3.3 | -2.2±3.1 | -2.1±3.7 | 0.962 |
| Change in NEWS score from baseline day 14 | -2.2±3.2 | -2.2±2.0 | -2.2±4.0 | 0.989 |
| Change in ROX index from baseline at day 7 | 5.6±6.6 | 4.9±6.6 | 6.6±6.5 | 0.257 |
| Change in ROX index from baseline at Day 14 | 7.2±5.3 | 6.5±5.8 | 7.8±4.8 | 0.537 |
| Secondary Infection* (%) | 17 (20.2) | 9 (17.6) | 8 (24.2) | 0.462 |

\*Secondary infection includes Ventilator associated pneumonia (VAP) and blood-stream infection (bacteremia).

A univariate analysis was then done to compare baseline characteristics and outcomes among living and deceased patients in both groups. The analysis revealed that patients who died were older (57.8 + 12.7 years versus 72.8+ 8.7 years, p<0.0001) and had more cardiac and renal comorbidities (Table-4). Additionally, patients who died had more severe disease at baseline indicated by significantly worse ROX index and NEWS scores and increased need for invasive mechanical ventilation (Table-5). Clinical Outcomes for patients in both groups are summarized in Table-6. Changes in each inflammatory marker post treatment initiation in both groups are represented in Figure-2. Interestingly, the amelioration of inflammatory markers such as CRP, fibrinogen and stabilization of lymphopenia were similar among the groups. However, compared to those who survived, patients who died had a significant worsening in their inflammatory markers (Figure-2).

**Table 4-.** Baseline Characteristics Comparing Living versus Deceased patients:

| Demographics | Total<br>n=84 | Living<br>n=68 | Deceased<br>n=16 | P-value |
| --- | --- | --- | --- | --- |
| Gender (%) |  |  |  | 0.094 |
| Male | 53 (63.1) | 40 (58.8) | 13 (81.3) |  |
| Female | 31 (36.9) | 28 (41.2) | 3 (18.8) |  |
| Mean Age, years | 60.6±13.4 | 57.8±12.7 | 72.8±8.7 | <0.0001 |
| Ethnicity (%) |  |  |  | 0.141 |
| AA | 45 (53.6) | 35 (51.5) | 10 (62.5) |  |
| Hispanic | 25 (29.8) | 22 (32.4) | 3 (18.8) |  |
| Caucasian | 13 (15.5) | 11 (16.2) | 2 (12.5) |  |
| Not known /NON<br>Hispanic | 1 (1.2) | 0 (0.0) | 1 (6.3) |  |
| Mean BMI<br>(kg/m<sup>2</sup>) | 31.4±8 | 32.6±8.0 | 26.7±5.7 | 0.008 |
| COVID PCR<br>positive status | 65 (77.4) | 51 (75.0) | 14 (87.5) | 0.282 |
| Therapy given: |  |  |  | 0.464 |
| Anakinra/IVIG | 51 (60.7) | 40 (58.8) | 11 (68.8) |  |
| Tocilizumab (%) | 33 (39.3) | 28 (41.2) | 5 (31.3) |  |
| **Comorbidities** |  |  |  |  |
| COPD (%) | 23 (27.4) | 17 (25.0) | 6 (37.5) | 0.313 |
| Asthma (%) | 8 (9.5) | 8 (11.8) | 0 (0.0) | 0.149 |
| HTN (%) | 60 (71.4) | 46 (67.6) | 14 (87.5) | 0.114 |
| DM (%) | 36 (42.9) | 29 (42.6) | 7 (43.8) | 0.936 |
| CAD (%) | 16 (19.0) | 8 (11.8) | 8 (50.0) | 0.0005 |
| CHF (%) | 17 (20.2) | 11 (16.2) | 6 (37.5) | 0.056 |
| CKD (%) | 12 (14.3) | 7 (10.3) | 5 (31.3) | 0.031 |
| ESRD on dialysis (%) | 11 (13.1) | 5 (7.4) | 6 (37.5) | 0.001 |
| Malignancy (%) | 4 (4.8) | 2 (2.9) | 2 (12.5) | 0.106 |
| HIV (%) | 4 (4.8) | 4 (5.9) | 0 (0.0) | 0.320 |
| Highest Corticosteroid dose (Methylprednisolone) (mg) | 229.1±142.1 | 217.8±140.6 | 277.2±142.6 | 0.133 |

**Table 5-.** Baseline Laboratory, Respiratory Function and Clinical Data in Living versus Deceased Patients

|  | **Total**<br>**n=84** | **Living**<br>**n=68** | **Deceased**<br>**n=16** | **P value** |
| --- | --- | --- | --- | --- |
| **Baseline laboratory Inflammatory Markers** |  |  |  |  |
| CRP (mg/L) | 8.9±7.0 | 8.4±6.3 | 10.6±9.2 | 0.288 |
| Ferritin (ng/mL) |  |  |  | 0.097 |
| Mean (SD) | 1599.2±5084.2 | 581.0±516.4 | 5672.2±10664.8 |  |
| Median (IQR) | 563.5 (247.0-993.0) | 468.5 (202.0-821.5) | 1359.5 (648.0-2610.0) |  |
| LDH (U/L) | 446.4±463.4 | 357.9±194.9 | 794.6±894.1 | 0.081 |
| AST (U/L) |  |  |  | 0.175 |
| Mean (SD) | 116.8±386.5 | 55.8±92.4 | 356.9±816.1 |  |
| Median (IQR) | 37.5 (25.0-66.0) | 35.0 (23.0-48.0) | 71.0 (36.0-217.0) |  |
| ALT (U/L) |  |  |  | 0.240 |
| Mean (SD) | 97.1±250.5 | 64.4±107.2 | 226.0±507.3 |  |
| Median (IQR) | 43.0 (22.0-69.0) | 43.0 (23.0-61.0) | 39.0 (17.0-134.0) |  |
| Albumin (g/dL) | 2.6±0.4 | 2.6±0.4 | 2.4±0.4 | 0.132 |
| D-dimer (ng/mL) |  |  |  | 0.170 |
| Mean (SD) | 4328.3±14055.6 | 2065.7±4251.7 | 13540.1±29531.0 |  |
| Median (IQR) | 843.0 (521.0-1745.0) | 814.0 (514.0-1291.0) | 1642.5 (830.0-7498.0) |  |
| Fibrinogen (mg/mL) | 519.3±163.7 | 547.7±143.0 | 421.1±196.1 | 0.007 |
| Triglycerides (mg/mL) | 141±87.8 | 139.5±87.1 | 145.8±93.1 | 0.810 |
| Absolute Lymphocyte Count (K/mm<sup>3</sup>) | 0.9±0.6 | 1.0±0.6 | 0.7±0.5 | 0.075 |
| BUN/Creatinine ratio | 23.9±12.4 | 23.9±10.0 | 23.8±19.8 | 0.978 |
| **Respiratory Function** |  |  |  |  |
| Mean ROX day 0 | 11.6±7.5 | 12.7±7.4 | 6.6±6.3 | 0.003 |
| Maximum Oxygen Therapy |  |  |  | <0.0001 |
| None (%) | 6 (7.1) | 5 (7.4) | 1 (6.3) |  |
| NC (%) | 38 (45.2) | 38 (55.9) | 0 (0.0) |  |
| HFNT (%) | 20 (23.8) | 15 (22.1) | 5 (31.3) |  |
| NIPPV (%) | 3 (3.6) | 3 (4.4) | 0 (0.0) |  |
| IMV (%) | 16 (19.0) | 6 (8.8) | 10 (62.5) |  |
| ECMO (%) | 1 (1.2) | 1 (1.5) | 0 (0.0) |  |
| **Clinical Data** |  |  |  |  |
| NEWS score Day 0 | 6.3±3.6 | 5.7±3.3 | 9.2±3.5 | 0.0003 |

\*NC- Nasal cannula, HFNT- High flow nasal Therapy, NIPPV- Non-invasive Positive Pressure Ventilation, IMV - invasive mechanical Ventilation, ECMO- Extracorporeal Membrane oxygenation, NEWS - National early warning Score

**Table 6-.** Clinical Outcomes in Living versus Decreased Patients

| **Outcome** | **Total**<br>**n=84** | **Living**<br>**n=68** | **Deceased**<br>**n=16** | **P-value** |
| --- | --- | --- | --- | --- |
| Intubation (%) | 16 (19.0) | 6 (8.8) | 10 (62.5) | <0.0001 |
| Need for ICU (%) | 36 (52.9) | 22 (42.3) | 14 (87.5) | 0.002 |
| Composite outcome (death, intubation, ICU admission) (%) | 38 (45.2) | 22 (32.4) | 16 (100) | <0.0001 |
| Hospital Length of Stay (LOS) (days) | 14±10.4 | 13.8±10.9 | 14.6±8.1 | 0.783 |
| ICU LOS (days) |  |  |  | 0.974 |
| Mean | 8.5±7.0 | 8.5±8.3 | 8.4±4.6 |  |
| Median (IQR) | 6.5 (3.0-12.0) | 5.0 (3.0-12.0) | 8.5 (6.0-12) |  |
| Duration of Invasive Mechanical of Ventilation (days) | 13.3±10.9 | 15.1±10.3 | 11.8±11.8 | 0.559 |
| Change in NEWS score from baseline at Day 7 | -2.1±3.3 | -2.5±3.1 | -0.7±3.7 | 0.050 |
| Change in NEWS score from baseline at day 14 | -2.2±3.2 | -2.5±2.9 | -0.8±4.1 | 0.274 |
| Change in ROX index from | 5.6±6.6 | 6.7±6.5 | 0.8±4.5 | 0.0010 |
| baseline at day 7 |  |  |  |  |
| Change in ROX<br>index from<br>baseline at Day 14 | 7.2±5.3 | 7.7±5.0 | 4.7±6.3 | 0.245 |
| Secondary<br>Infection* (%) | 17 (20.2) | 10 (14.7) | 7 (43.8) | 0.009 |

\*Secondary infection includes Ventilator associated pneumonia (VAP) and blood-stream infection (bacteremia).

**Figure 2.**
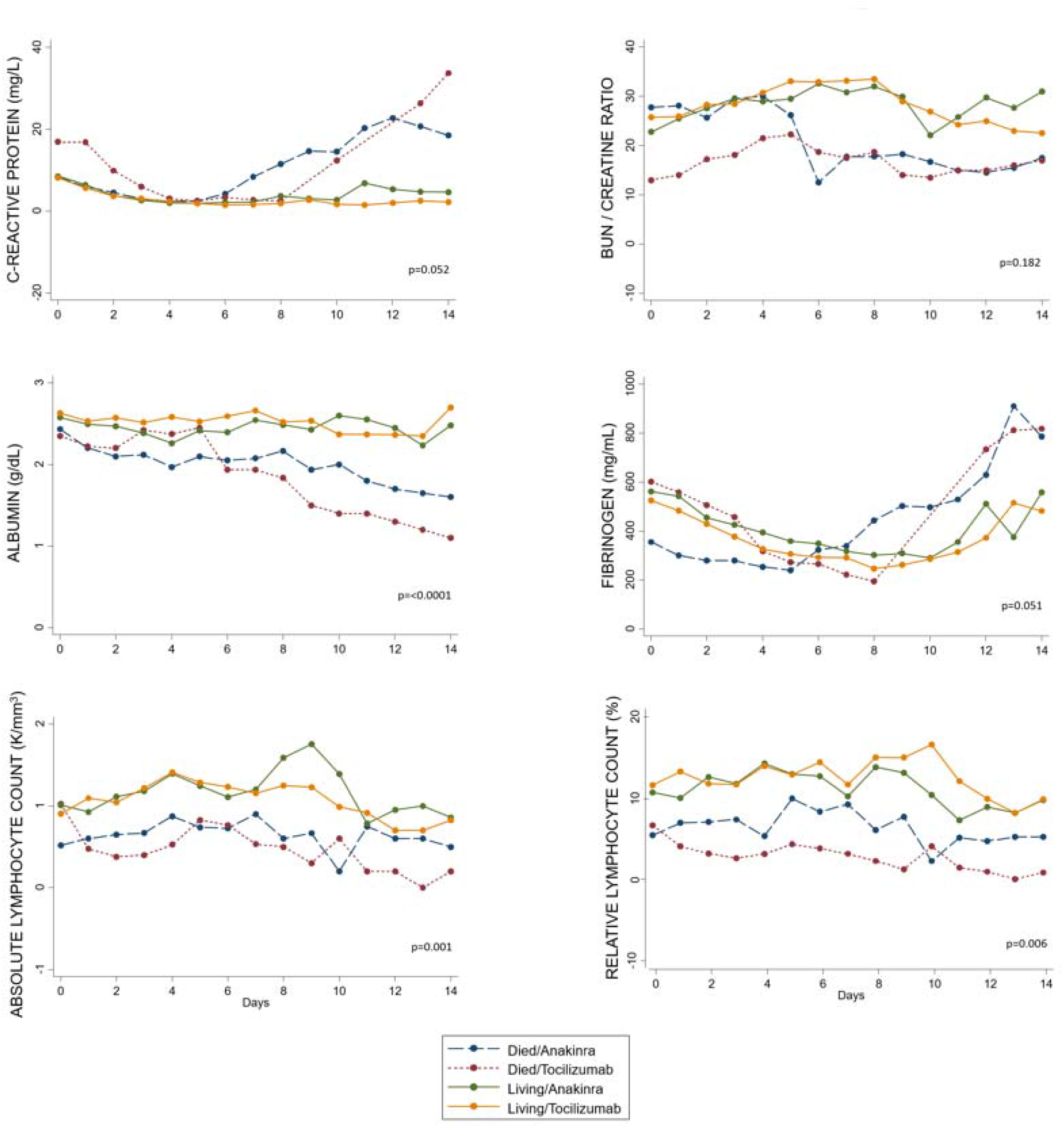
Comparison of the mean data of each inflammatory marker concentration over time among patients of the Anakinra/IVIG and Tocilizumab groups. Day 0 corresponds to the date of initiation of therapy. Data are presented for patients who survived and those who died in each treatment group on separate curves for comparison. Displayed within each graph is the p value for the rate of change for each inflammatory marker comparing living and deceased at day 14 post initiation of therapy.

## Discussion

To our knowledge, this is the first study that compares two unique treatments for COVID-19 pneumonia in patients with hyperinflammatory response. Although their mechanisms are different, the desired outcomes appear to be somewhat similar. As noted, at baseline, both patient populations are comparable in terms of demographics and comorbidities. They also have similar inflammatory responses indicated by elevated levels of CRP, LDH, Fibrinogen and D-dimers in both groups. However, patients who received Tocilizumab were sicker in regard to their clinical data (lower baseline ROX index and higher NEWS score). To some extent, this reflects a selection bias as intravenous biologic treatment was preferred over subcutaneous therapies to avoid delay in treatment response in rapidly deteriorating patients. Subcutaneous injections can have variable absorption characteristics, particularly with factors such as obesity and anasarca that can extend the half-life, and do not achieve the maximum serum concentrations possible with intravenous administration. These issues led us to add intravenous IgG to the anakinra regimen so that a concomitant faster treatment could be provided. Nevertheless, the outcomes were similar in both groups in regard to mortality, intubation, ICU admission, length of stay and improvement in inflammatory markers. There was also no significant difference in the rate of secondary infection in both groups.

Based on our results, it appears that regardless of the advanced therapy given (Anakinra/IVIG or Tocilizumab), patients who are older, who have cardiac or renal comorbidities, and show more disease burden and organ dysfunction (indicated by ROX index and NEWS scores) are at highest risk for mortality. Another important finding in our study is that response to therapy at 7- and 14-days may predict overall outcomes for the patient. Baseline inflammatory markers did not differentiate those who died from the ones who survived; however, failure of those markers to improve after therapy appears to have important prognostic value. Indeed, ferritin levels that fail to decline in classical HLH also correlate with worse outcomes.

Given the retrospective design, it may be challenging to make a comparison and draw conclusions between our study, and the studies that have been published recently in the literature. However, we believe that our results are fairly similar in regard to mortality and safety outcomes. In studies that evaluated Anakinra as a treatment for COVID-19 pneumonia, mortality ranged anywhere between 10 to 25%, (21.6% in our population).^19,20^ Improvement in clinical outcomes was also noted in several retrospective, observational studies, case series and case reports. These outcomes include ICU admission, improvement in inflammatory markers, radiographic findings and respiratory parameters.^14,19–25^

It is important to mention that Anakinra was used subcutaneously in many of the aforementioned studies, and the dose used was smaller than the one we considered in our practice.^19,22–25^ We argue that this should take into account the onset of action of the medication, and thus perhaps higher doses of Anakinra to achieve the desired rapid effectiveness are required, especially in treating critically ill patients. This may not be the case of concern when using intravenous Anakinra, however this form may not be widely available. In fact, in a study by Cavalli et al. high dose intravenous Anakinra achieved better outcomes compared to lower dose treatment or standard of care. In the same study, low dose subcutaneous Anakinra was initially used but the treatment was stopped due to paucity of effect after 7 days. Similarly, a case series by Millan et al. revealed that patients who benefitted from receiving subcutaneous Anakinra were those who received it early in the course of disease (≤36 hours). In our study, we chose to give a high dose of subcutaneous Anakinra given lack of availability of the intravenous form; in addition, we used IVIG concomitantly in the first three days to augment the action of Anakinra, similar to other cytokine storm protocols.^13-15^

Our data on treating COVID-19 patients with tocilizumab are similar to those treated with Anakinra favoring this advanced therapy, and mostly align with the limited retrospective reports and case series that have been published thus far. Mortality ranged between 16 to 52% (15.2% in our study) with higher mortality observed in older, sicker patients, severe ARDS and especially in those who were on invasive mechanical ventilation.^26–32^. Secondary infection, which may be a concern when using IL-1 or IL-6 blockers were not significantly different between the two therapies. Our results on superinfection are similar to what have been reported in the literature, 12 to 14% with higher rates of superinfections observed in mechanically ventilated and ICU patients.^20,26,28,30^ Likewise, the limited HLH literature does not suggest a substantially increased risk of bacterial superinfection despite combination anti-inflammatory therapy, with anakinra potentially less myelotoxic than anti-interleukin 6 therapy.^13,15^

Interestingly, despite having two different mechanisms, both treatments resulted in a similar rate of improvement in clinical data and inflammatory markers. This suggests a common pathway of these two approaches that results in amelioration of inflammation parameters and eventually controlling the storm. This is extremely important in patients with severe respiratory failure, which may correlate to the decrease in CD4 and NK cells, the reduction in cytotoxic potentials, and the increase in proinflammatory cytokines levels, such as IL-1, IL-6, tumor necrosis factor alpha, and interferon gamma. In these patients Tocilizumab has been shown to restore IL-6 mediated response of CD4 and NK cells and increase the number of circulating lymphocytes.^10, 12, 33^ Anakinra instead reduces circulating levels of the IL-1β and is thought to help in halting the inflammatory cascade caused by SARS-CoV2 virus.

Corticosteroids are added to also blunt the inflammatory response. At our institution, we initiated corticosteroids in patients with moderate-severe disease burden based on experience from colleagues in China.^34^ One study has suggested that adding methylprednisolone to Tocilizumab decreases the risk of death.^35^ Results of randomized data from >6000 patients in the United Kingdom suggest a mortality benefit with the use of dexamethasone.^7^

The main limitation of our study is due to its retrospective, single center observational design. This design may put our study at risk for possible selection bias given no randomization to treatment or standard of care arm. Furthermore, patients were selected for each therapy depending on drug availability in our institution. The fact that we found equivalent outcomes between the two groups does provide encouragement in this regard, as real-world situations may limit the choice of drug availability at different institutions. The ubiquitous use of steroids at our institution might present a confounding factor, however anti-interleukin therapy was often added after failure of corticosteroid treatment.

## Conclusion

In summary, our study suggests that treating COVID-19 pneumonia associated with cytokine storm features with either subcutaneous Anakinra/IVIG or intravenous Tocilizumab is associated with improved clinical outcomes in most subjects. The choice of therapy does not seem to affect morbidity or mortality. We believe that the addition of IVIG may offset the less efficacious use of subcutaneous anakinra, especially in centers where the intravenous form is not available. Our data provide meaningful information to clinicians that care for patients with COVID-19 disease. Randomized, prospective clinical trials are needed to confirm our study findings.

## Data Availability

All data related to patients information are stored on a password-protected file on Temple University secure computers. In addition, the data is de-identified to protect patients information. The data will be available upon request for any verification purposes.

## Supplement

**Table-7.** P values comparing the rate of change in inflammatory markers at day 14 compared to baseline

| Inflammatory Marker | p value comparing living and deceased at day 14 | p value comparing Anakinra and Tocilizumab group at day 14 |
| --- | --- | --- |
| CRP | 0.052 | 0.995 |
| BUN/Creatinine ratio | 0.182 | 0.193 |
| Albumin | <0.0001 | 0.468 |
| Fibrinogen | 0.051 | 0.586 |
| Absolute lymphocyte count | 0.001 | 0.299 |
| Relative lymphocyte count | 0.006 | 0.967 |

